# Detection of brain somatic variation in epilepsy-associated developmental lesions

**DOI:** 10.1101/2021.12.06.21267079

**Authors:** Tracy A. Bedrosian, Katherine E. Miller, Olivia E. Grischow, Hyojung Yoon, Kathleen M. Schieffer, Stephanie LaHaye, Anthony R. Miller, Jason Navarro, Jesse Westfall, Kristen Leraas, Samantha Choi, Rachel Williamson, James Fitch, Kristy Lee, Sean McGrath, Catherine E. Cottrell, Vincent Magrini, Jeffrey Leonard, Jonathan Pindrik, Ammar Shaikhouni, Daniel R. Boué, Diana L. Thomas, Christopher R. Pierson, Richard K. Wilson, Adam P. Ostendorf, Elaine R. Mardis, Daniel C. Koboldt

## Abstract

Epilepsy-associated developmental lesions, including malformations of cortical development and low-grade developmental tumors, represent a major cause of drug-resistant seizures requiring surgical intervention in children. Brain-restricted somatic mosaicism has been implicated in the genetic etiology of these lesions; however, many contributory genes remain unidentified. We enrolled 50 children undergoing epilepsy surgery into a translational research study. We performed exome and RNA-sequencing of resected brain tissue samples to identify somatic variation. We uncovered candidate disease-causing somatic variation affecting 28 patients (56%), as well as candidate germline variants affecting 4 patients (8%). We confirmed somatic findings using high-depth targeted DNA sequencing. In agreement with previous studies, we identified somatic variation affecting SLC35A2 and MTOR pathway genes in patients with focal cortical dysplasia. Somatic gains of chromosome 1q were detected in 30% (3 of 10) Type I FCD patients. Somatic variation of MAPK pathway genes (i.e., *FGFR1, FGFR2, BRAF, KRAS*) was associated with low-grade epilepsy-associated developmental tumors. Somatic structural variation accounted for over one-half of epilepsy-associated tumor diagnoses. Sampling across multiple anatomic regions revealed that somatic variant allele fractions vary widely within epileptogenic tissue. Finally, we identified putative disease-causing variants in genes (*EEF2, NAV2, PTPN11*) not yet associated with focal cortical dysplasia. These results further elucidate the genetic basis of structural brain abnormalities leading to focal epilepsy in children and point to new candidate disease genes.

## Introduction

Early-onset focal epilepsy is commonly associated with developmental brain lesions, including focal cortical dysplasia (FCD) and long-term epilepsy-associated tumors (LEATs). These lesions are among the most common etiologies encountered during surgical intervention for epilepsy, as their associated seizures tend to be resistant to anti-seizure medications^1^. Cases of FCD are broadly classified by the International League Against Epilepsy (ILAE) into three categories based on histological subtype^2^. Type I FCD is an isolated lesion characterized by focal abnormalities of radial (Type Ia), tangential (Type Ib), or radial and tangential (Type Ic) cortical lamination. Type II FCD is characterized by dysmorphic neurons either alone (Type IIa) or in the presence of balloon cells (Type IIb). Type III FCD reflects cortical dyslamination associated with another type of lesion, specifically hippocampal sclerosis (Type IIIa), glial or glioneuronal tumors (Type IIIb), vascular malformations (Type IIIc), or other early-life lesions including trauma, injury, or encephalitis (Type IIId). Likewise, LEATs represent a histologically heterogeneous group of entities. These low-grade tumors often present with early-onset drug-resistant epilepsy as the primary clinical sign^3^. Ganglioglioma (GG), dysembryoplastic neuroepithelial tumor (DNET), low-grade glioneuronal tumors, pleomorphic xanthoastrocytoma (PXA), and polymorphous low-grade neuroepithelial tumor of the young (PLNTY) have been described as LEATs, although no consensus definition is available from the World Health Organization (WHO)^4^.

Somatic variation in brain tissue has long been proposed as a genetic mechanism for FCD and LEATs given their focal and developmental nature. Over the past decade, advances in genome sequencing technology, along with the availability of affected brain tissue upon surgical treatment, have enabled studies to elucidate the genetic basis of FCD and LEATs. Indeed, somatic mutations affecting a mosaic population of brain cells have been implicated in both lesion types. Recent cohort analyses revealed somatic variation of *SLC35A2* associated with up to one-third of Type I FCD cases and somatic variation of PI3K-AKT-MTOR pathway genes in approximately half of Type II FCD cases^5-8^. Somatic alterations of the RAS-RAF-MAPK pathway and PI3K-AKT-MTOR pathways have been associated with LEATs, with somatic *BRAF* p.V600E mutations accounting for more than half of ganglioglioma cases and somatic *FGFR1* variants found in more than half of DNETs^9,10^. Importantly, understanding the molecular underpinnings of epilepsy-associated lesions may pave the way for the development of targeted therapies.

Despite current progress, a large proportion of molecularly characterized FCD cases do not have an apparent genetic etiology. Here we present an approach to detect somatic variation in epilepsy-associated developmental lesions. Fifty patients undergoing surgical resection of FCD- or LEAT-affected tissue were characterized in terms of clinical, neuropathological, and genetic findings. For each patient, multiple samples of affected tissue when available were analyzed by high-depth exome sequencing and RNA-sequencing to detect somatic variation across the entire protein-coding genome. This approach yielded somatic findings in previously described FCD- or LEAT-associated genes, as well as novel candidate genes.

## Materials and Methods

### Patient enrollment and sample collection

Patients undergoing surgical treatment for drug-resistant epilepsy at Nationwide Children’s Hospital were enrolled on an IRB-approved research protocol (Supplementary Table 1). Blood and snap frozen brain tissue samples (range: 1-13 samples per patient, mean: 3.2 samples) were collected at the time of surgical resection for genomic analysis. DNA and RNA were co-extracted from each sample. Formalin fixed paraffin embedded tissue blocks were prepared from all tissues and submitted for standard clinical neuropathologic evaluation.

### Variant detection by exome sequencing

DNA isolated from blood and brain tissue was prepared for enhanced exome sequencing using NEBNext Ultra II FS DNA Library Prep Kit (New England Biolabs, Ipswich, MA). IDT xGen Exome Research Panel v1.0 enhanced with the xGenCNV Backbone Panel-Tech Access (Integrated DNA Technologies, Coralville, IA) was used for target enrichment by hybrid capture. Final libraries were sequenced on an Illumina HiSeq 4000 or NovaSeq 6000 to generate paired-end 151-bp reads. A custom pipeline was used for alignment to human reference genome build GRCh37 and secondary analysis^11^. For candidate disease-causing germline variants, parental testing of salivary DNA (when available) via sanger sequencing was performed to determine the mode of inheritance.

Germline variants were called by GATK v.4.0.5.1 HaplotypeCaller and somatic variants were called by MuTect-2^12,13^. Germline variants were filtered based on the following features: GATK quality score (≥ 30), population frequency (≤ 1%), depth of sequencing (≥ 8 reads), variant allele fraction (VAF; ≥ 20%), protein-coding region or distance from canonical splice site ≤ 3 bp, damaging prediction scores (CADD ≥ 15; SIFT ≤ 0.05; GERP++ ≥ 2), and relevance of gene-disease association with patient phenotype using OMIM and ClinVar databases. We focused on 2,897 genes present on an in-house curated list of epilepsy-related genes pulled from commercial genetic testing panels and epilepsy literature (Supplementary Data File). Passing germline candidate variants were assessed by ACMG/AMP criteria (Supplementary Table 2)^14^.

Somatic variants were filtered based on the following features: MuTect-2 = “PASS”, GATK quality score (≥ 30), protein-coding region or distance from canonical splice site ≤ 3 bp, depth of sequencing (≥ 8 total reads), absence from blood comparator, alternate allele reads (≥ 4), VAF (≥ 1%), and presence of gene on an in-house curated epilepsy gene list (Supplementary Data File). Passing variants were reviewed in Integrated Genomics Viewer (IGV) to assess for strand bias, read-end bias, or other biases which may suggest sequencing artifacts. A full list of passing somatic variants is presented in Supplementary Table 3. Copy number variation was called using VarScan2 using exome sequencing data from blood as a comparator for each brain tissue sample^15^.

### Targeted sequencing validation

Somatic variants were validated by targeted sequencing, as previously described, with exception of *PTEN* variants which were validated by a targeted capture method, as previously described^16,17^. Human reference DNA GM24143 or HG04217 (Coriell Institute) was used as a negative control. Primer sequences for targeted amplification are provided in Supplementary Table 4. SAMtools mpileup and mpileup2cns VarScan (version 2.3.4) commands were used to generate read counts for variants of interest from raw, deduplicated BAM files.

### RNA-sequencing and analysis

RNA isolated from brain tissue was treated with DNase and ribodepletion prior to constructing libraries using NEBNext Ultra II Directional RNA Library Prep Kit. Paired-end 151-bp reads were generated on an Illumina HiSeq 4000 and then low-quality reads (q<10) and adaptor sequences were eliminated from raw reads using bbduk version 37.64. Reads were aligned to the GRCh38.p9 assembly of the human reference genome from NCBI using version 2.6.0c of the aligner STAR^18^. Fusions were detected in LEAT cases using an ensemble approach of seven fusion calling algorithms as described previously^19^. Fusions identified by at least three fusion callers were reviewed for biological relevance, including literature searches of the gene functions and previous association to disease. Internal tandem duplication events were called by CICERO using default parameters and were reviewed manually in IGV for authenticity and reviewed for biological relevance based on gene function^20^.

## Results

### Study design and patient cohort

Fifty patients (ages 0-21 years) undergoing surgical treatment for drug-resistant epilepsy at Nationwide Children’s Hospital were enrolled on a research protocol (Supplementary Table 1). Each patient was evaluated by a multidisciplinary pediatric clinical care team for neuroimaging, electrophysiology, and post-surgical neuropathology prior to genomic analysis (Figure 1A). Patients included in this study were diagnosed with FCD and/or a LEAT (Figure 1B), including ganglioglioma (GG), dysembryoplastic neuroepithelial tumor (DNET), pleomorphic xanthoastrocytoma (PXA), polymorphous low-grade neuroepithelial tumor of the young (PLNTY), or diffuse low-grade glioma MAPK pathway-altered, a new tumor entity included in the 2021 World Health Organization classification of tumors of the central nervous system^21^. Notably, six LEAT cases were initially diagnosed as low-grade glioneuronal tumors not elsewhere classified per cIMPACT-NOW guidelines, but received a refined diagnosis based on genomic findings^22^. Specific subtyping of neuropathological diagnoses is shown in Figure 1C. The age of seizure onset varied by diagnosis with FCD patients presenting earlier than LEAT patients on average (two-tailed t-test, P=0.001; Figure 1D). The median age at surgery across all diagnostic groups was 11 years (range: <1 month – 21 years).

**Figure 1:**
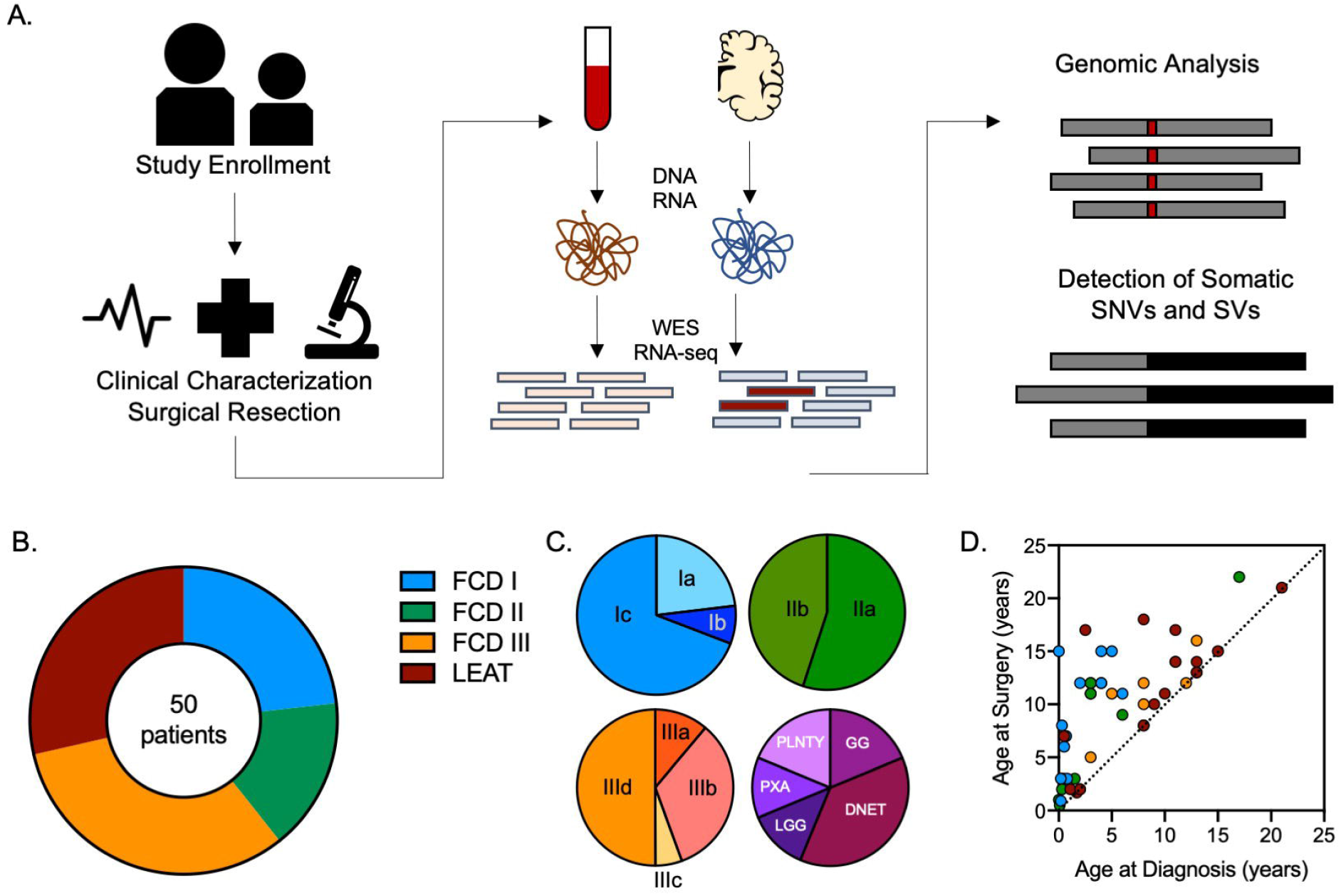
Study design and patient cohort. **(A)** Overview of study design and workflow. **(B)** Cohort composition by general neuropathology diagnosis. Six cases of tumor with surrounding dysplasia are counted in both LEAT and FCD IIIb classes. **(C)** Specific neuropathology observed (N=3 FCD Ia, N=1 FCD Ib, N=9 FCD Ic; N=5 FCD IIa, N=4 FCD IIb; N=2 FCD IIIa, N=6 FCD IIIb, N=1 FCD IIIc, N=9 FCD IIId; N=6 DNET, N=3 GG, N=3 PLNTY, N=2 PXA, N=2 LGG). **(D)** Age of surgery by age of diagnosis for each patient.

### Known and novel somatic variation in focal epilepsy

To identify candidate genetic variation underlying each patient’s diagnosis, we performed exome sequencing (∼200x coverage) of matched blood and resected brain tissue samples (Figure 2A). Multiple anatomic regions were sequenced for a given patient when available (Supplementary Table 2). We identified putative disease-causing somatic variation in 28 patients (56%) and candidate germline variants in 4 patients (8%) (Figure 2B and Supplementary Table 2). Notably, 100% of LEAT cases had a genetic finding, but a substantial fraction of FCD cases had no apparent genetic finding, ranging from over half of Type II FCD cases to less than a quarter of Type I FCD cases. Some of the somatic variation that we identified in this cohort has been described in association with FCD and LEATs, including *MTOR, RHEB, PTEN*, chromosome 1q gain, *SLC35A2, BRAF, FGFR1*, and *FGFR2* (Figure 2C). However, we also identified novel candidate genes (*EEF2, NAV2*, and *PTPN11*) not previously associated with FCD (discussed below). We validated somatic variants by high-depth targeted amplicon sequencing (69,000x coverage on average; Supplementary Table 3). For variants detected above 2% VAF in exome sequencing analysis, the VAFs were highly concordant between targeted and exome sequencing (Figure 2D). Variants detected at less than 2% VAF in exome analysis were typically refuted by targeted validation (80% of examples; Figure 2D inset). Interestingly, somatic variant allele fractions varied widely depending on the anatomic region sequenced, highlighting sampling bias as a potential obstacle to variant detection in somatic mosaic diseases (Figure 2E). For example, in one case of Type I FCD, a causal somatic variant in *SLC35A2* was detected at >13% targeted sequencing VAF in hippocampus, but <2% VAF in the occipital lobe, as illustrated by blue shading in Figure 2E. Sequencing multiple affected regions where possible may provide a greater opportunity for variant detection.

**Figure 2:**
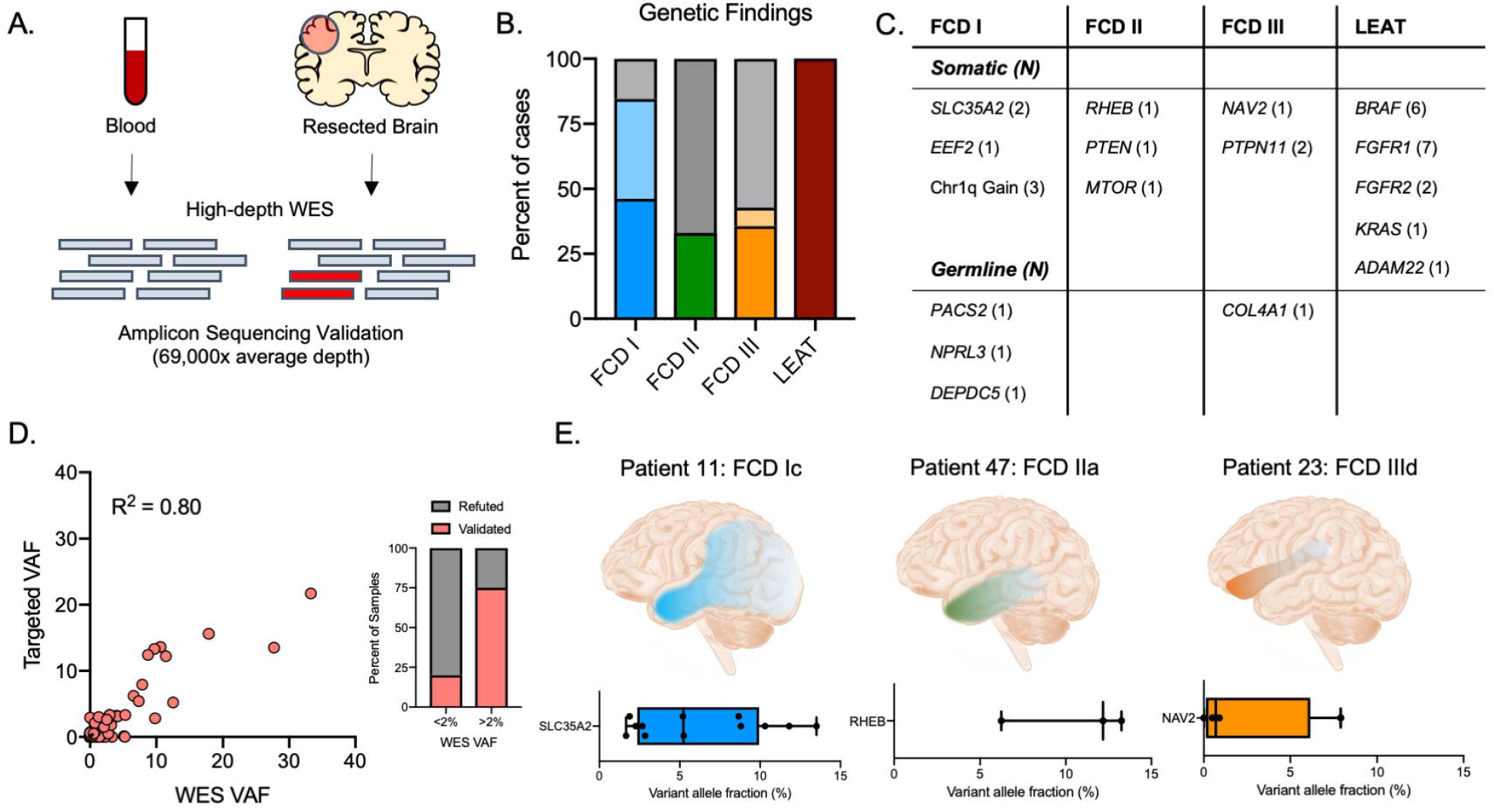
Genetic findings by exome sequencing. **(A)** Overview of DNA sequencing strategy. **(B)** Proportion of cases with a candidate causal genetic finding with somatic findings represented by dark shading and germline findings represented by light shading. Gray shading indicates cases with no genetic finding. **(C)** Summary of genes with candidate causal variants identified in each pathology and number (N) of patients affected. **(D)** Variant allele fractions from exome sequencing versus targeted validation sequencing for each validated variant/sample. Inset shows percent of samples validated by targeted sequencing when initially detected at WES VAF less than or greater than 1%. **(E)** Example distributions of candidate somatic variants across tissue samples within individual patients. Shading of illustrations represents approximate anatomical depictions of the VAFs from targeted sequencing validation shown below.

### Novel candidate genes associated with FCD

We identified putative disease-causing somatic variants in three genes not previously associated with FCD. First, we identified a somatic missense variant in *EEF2* (NM_001961.3:c.1132G>A:p.Asp378Asn) at 4% VAF in a Type I FCD patient. This variant is absent from the gnomAD population database and predicted to be damaging by 17/27 computational tools reported by Varsome^23^. *EEF2* is implicated in translation initiation and elongation as part of the MTOR pathway. Both *in vitro* and *in vivo* functional studies support its role in GABAergic signaling and seizure susceptibility, making it a possible candidate for the patient’s phenotype^24,25^. Second, we detected a somatic missense variant in *NAV2* (NM_182964.5:c.1093G>A:p.Val365Met) in a Type II FCD patient at 8% variant allele fraction. This variant was present in only one individual in the gnomAD database^26^. *NAV2* has been implicated in neuronal migration and cerebellar development using model organisms^27,28^. Finally, in a patient with Type IIIa FCD, we found a somatic missense variant in *PTPN11* (NM_002834.5:c.1502G>A:p.Arg501Lys), which has been previously reported as Pathogenic in ClinVar in a germline setting among patients with Noonan syndrome. Our patient’s lesion was more diffuse than typical, encompassing regions of frontal, temporal, and parietal cortex in conjunction with hippocampal sclerosis. We observed the *PTPN11* variant in both sampled regions of cortex (frontal and parietal areas) at up to 3% VAF. We identified a second *PTPN11* missense variant (NM_002834.5: c.50A>G:p.Glu17Gly) ranging from 0-7.34% VAF in a different Type IIId FCD patient with a history of intraventricular hemorrhage. This variant is absent from the gnomAD database and predicted to damaging by 27/27 algorithms on Varsome. *PTPN11* has been described in relation to glioneuronal tumors, particularly in the context of Noonan syndrome, and implicated in hypothalamic hamartoma, but to our knowledge no study has identified somatic variation of *PTPN11* in FCD^29,30^. Interestingly, a mouse model of *PTPN11* deficiency in radial glial cells leads to developmental abnormalities of the cortex^31^. Future functional studies may be warranted to assess the effects of these variants in brain development. Notably, four additional genes were affected by validated somatic variants that we did not consider likely causal based on population frequency, lack of disease association, or presence of another more compelling candidate variant in the case (i.e., *POLR1A, SLC26A1, ENPP1*, and *ROCK2*; Supplementary Table 3).

### Structural variation in low-grade epilepsy-associated tumors

Complex structural rearrangements or fusion transcripts were detected by RNA-sequencing analysis in 9 LEAT cases, accounting for 69% of LEAT diagnoses overall (Figure 3A-B). All five cases of DNETs had evidence of internal tandem duplications within the *FGFR1* gene, all affecting the same general region of the gene (between exons 9 or 10 to exon 18; Figure 3C). Activating *FGFR1* mutations are a common molecular finding among DNETs, although the mechanistic link to epilepsy is not completely defined. Interestingly, one DNET with an *FGFR1* alteration also had a somatic frameshift variant affecting *ADAM22*, which has been previously associated with epilepsy^32^. The variant is absent from public databases and predicted to truncate most of the encoded protein. Four LEAT cases had evidence of fusion transcripts implicating either *FGFR1* or *FGFR2* with a variety of fusion partners, highlighting structural rearrangements as one genetic etiology of LEATs (Figure 3D). *BRAF* p.V600E mutations accounted for most remaining LEAT cases in our cohort.

**Figure 3:**
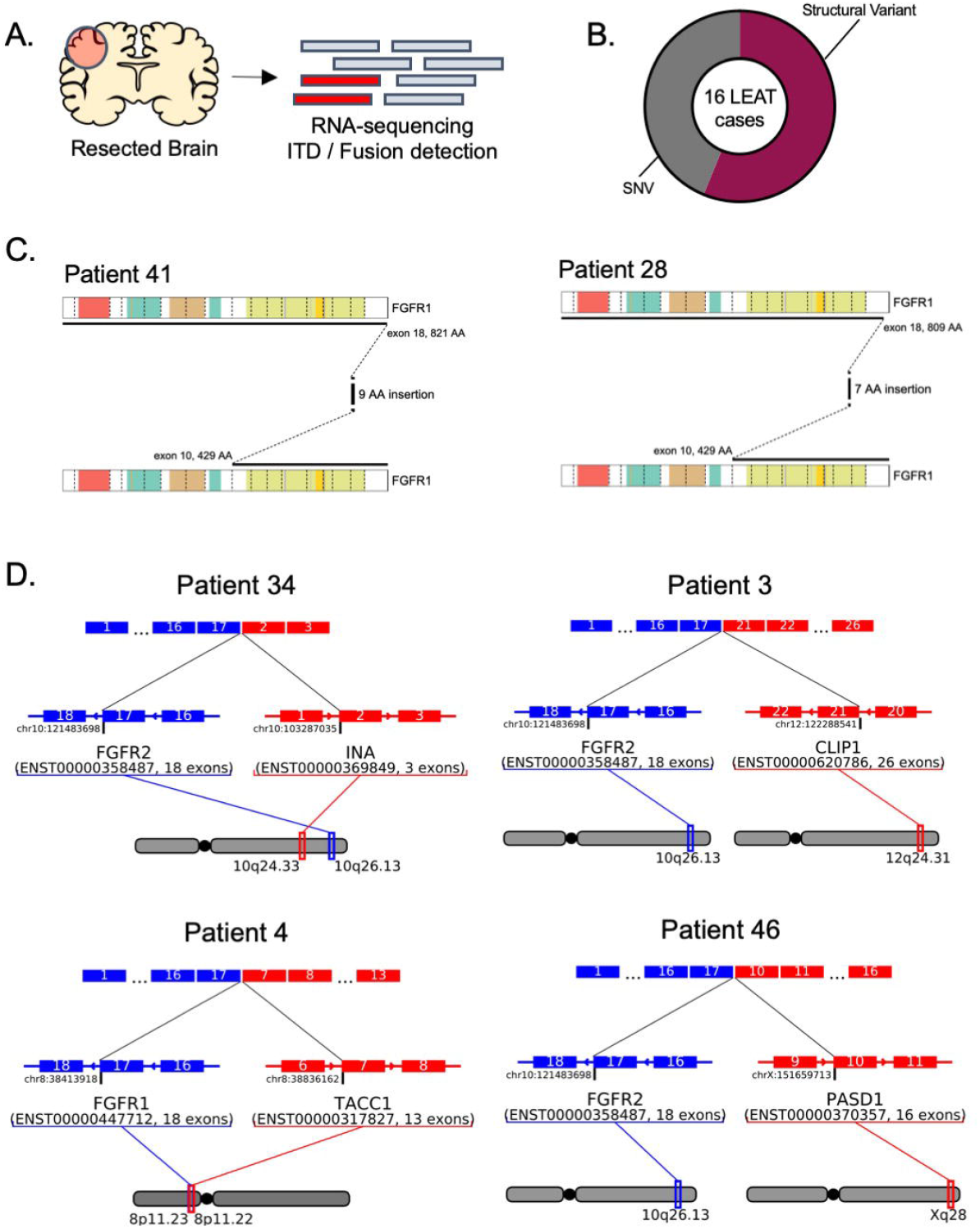
Somatic structural variation among LEATs. **(A)** Overview of RNA-sequencing strategy for structural variant detection. **(B)** Proportion of structural variants versus single-nucleotide variants among LEAT cases. **(C)** Two examples of *FGFR1* internal tandem duplications identified in this study. **(D)** Graphical summary of fusion transcripts identified in this study.

### Cortical dysplasia adjacent to tumors

Type IIIb FCD is characterized by cortical dysplasia surrounding a developmental tumor. In such cases, it is unclear whether the dysplasia and tumor represent distinct pathologies or whether they have the same etiology. In four cases with cortical dysplasia adjacent to a tumor, we independently analyzed samples containing only resected tumor or only cortical dysplasia for the presence of somatic variants. In all four cases, the causal variants were identified in the tumor, but not in the surrounding dysplastic tissue (Supplementary Table 3). This suggests that the dysplasia in these cases may have a different etiology or arise from non-cell autonomous effects, although it’s also possible that variants are present at levels below our detection limit.

### Rare germline variation in focal epilepsy

In addition to somatic variation, we identified four candidate disease-causing germline variants that were not detected during clinical characterization (Supplementary Table 2). We assessed a nonsense variant detected in *DEPDC5* as pathogenic according to ACMG/AMP variant interpretation criteria. *DEPDC5* mutations are known to be associated with familial focal epilepsy with variable foci^33^. In addition, we detected two variants of interest assessed as ‘likely pathogenic.’ First, we noted a *COL4A1* variant in a Type III FCD patient with a clinical history of intraventricular hemorrhage. *COL4A1* is associated with focal epilepsy and susceptibility to intracerebral hemorrhage^34^. Second, in a patient with FCD Type I, we identified a frameshift variant in *NPRL3*, a gene previously associated with autosomal dominant focal epilepsy^35^.

Finally, we noted a paternally inherited *PACS2* frameshift variant in a Type 1 FCD patient. *PACS2* has been associated with early-onset epilepsy and various central nervous system abnormalities but loss-of-function variants (as seen in our patient) are not a known mechanism of disease so we assessed it as a variant of unknown significance (VUS)^36^.

### Molecular correlates of somatic variation

Most of the somatic variation detected in FCD and LEAT cases was associated with the PI3K-AKT-MTOR pathway or MAPK pathway, respectively (Figure 4A). Interestingly, *PTPN11* (a.k.a. SHP2) has significant crosstalk among both MTOR and MAPK signaling pathways^37^. Somatic variants in *PTPN11* were detected in two (15%) of 12 Type III FCD patients. Type III FCD has been viewed as a highly heterogeneous group of pathologies, but this result suggests the possibility of underlying commonalities even among patients with distinct principal lesions. Somatic gains of chromosome 1q were detected in 3 of 10 Type I FCD cases (30%), suggesting that this may be a more common mechanism of FCD than previously appreciated (Figure 4B). The causal contributory genes associated with chromosome 1q gain remain unknown, but it is possible that MTOR pathway activation may be implicated given the presence of *AKT3* on chromosome 1q. Further, *SLC35A2* variants were identified in 2 of 10 Type I FCD cases (20%), which agrees with the findings of recent cohort analyses^5-7^. As *SLC35A2* plays a role in the glycosylation of many different proteins, its critical downstream effectors remain unspecified (Figure 4C).

**Figure 4:**
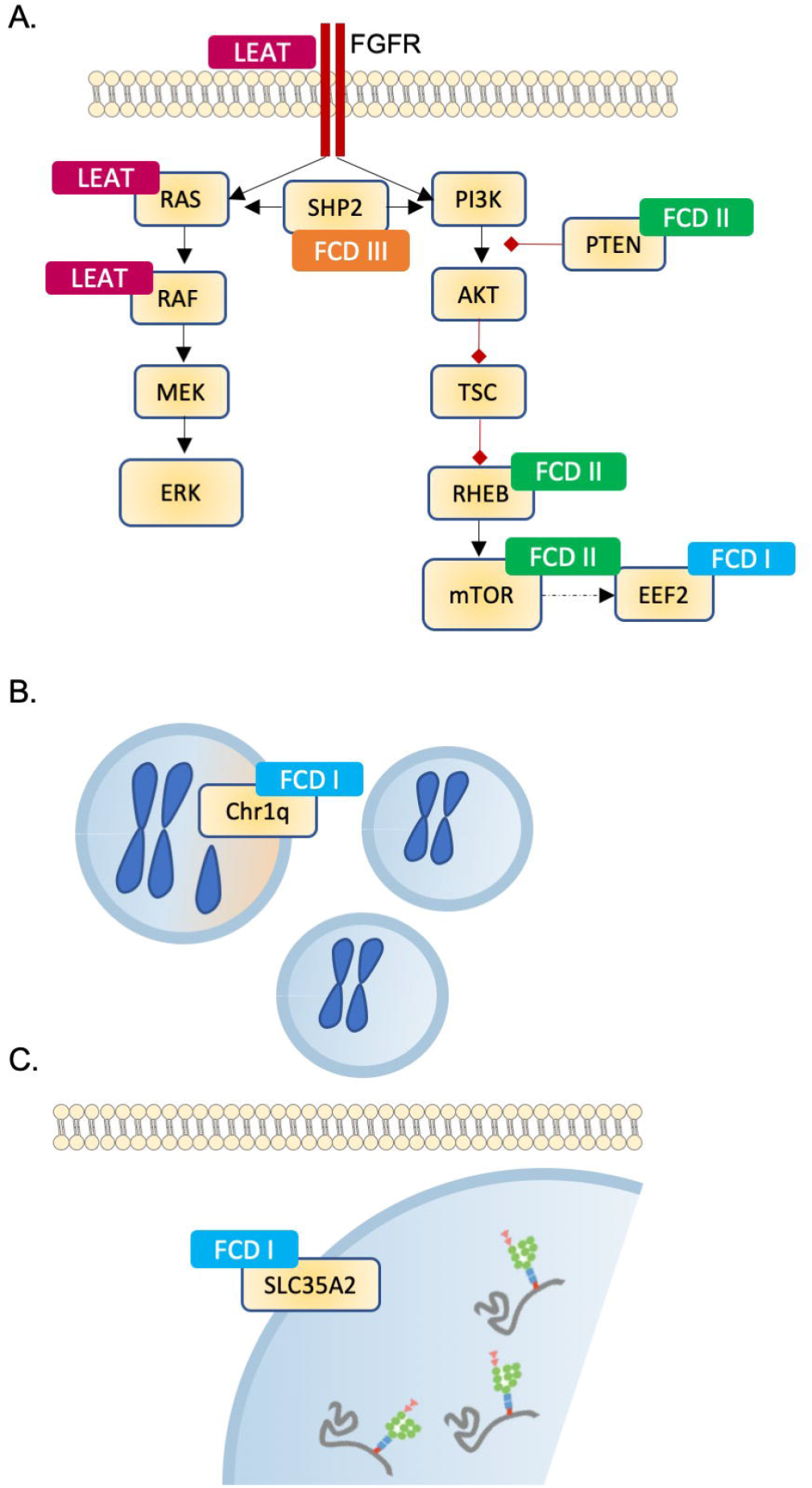
Molecular correlates of somatic variation. **(A)** Schematic of major molecular pathways implicated in FCDs and LEATs. **(B)** Chromosome 1q gain is a mechanism of Type I FCD, affecting 30% of patients in this cohort. **(C)** Glycosylation pathway disruption via *SLC35A2* is a molecular mechanism of Type I FCD.

## Discussion

Epilepsy-associated developmental lesions, including FCD and LEATs, are a major cause of drug-resistant epilepsy in children. Our results indicate that brain somatic variation is implicated in a high proportion of cases (approximately 40% of FCD cases and 100% of LEAT cases in this study). Nevertheless, a majority of FCD patients in our cohort remain without genetic diagnosis. This could be related to several factors, including limitations related to the specific tissue sampled. Interestingly, we observed that the burden of somatic variation within an individual brain may vary substantially, even within the epileptogenic region. Our results showed VAFs ranging from <1%-33% across brain samples within individual patients, highlighting the importance of tissue sampling to variant detection. In addition, depth of sequencing may be a limiting factor for somatic variants present at low allele fractions. However, our results suggest that variants detected at less than 2% variant allele fraction in exome sequencing data are unlikely to be validated by targeted deep sequencing and are likely artifacts. Finally, choice of methodology may affect success in variant detection. The addition of RNA-sequencing in our analyses allowed for the detection of structural variants that would not have been readily identified by exome sequencing analysis alone, leading to genetic findings in 100% of LEAT cases in this cohort.

Much of the somatic variation identified in this study implicates genes and pathways previously associated with focal epilepsy; however, three newly identified candidate genes (*EEF2, NAV2*, and *PTPN11*) may warrant future functional study. In particular, two of the Type III FCD patients (15%) harbor somatic variants in *PTPN11*, which has not been directly implicated in FCD to our knowledge, but certainly has a major role in brain development based on human and mouse data^30,31^. Although Type III FCD represents a heterogeneous mix of pathologies, there may be some common genetic mechanisms not yet fully appreciated. In addition, structural variation accounted for more than half of LEAT cases in this cohort, highlighting the presence of multiple forms of disease-causing somatic mosaicism within epilepsy-associated lesions, as well as the importance of screening for different kinds of somatic variation. Furthermore, by elucidating the genetic basis of several low-grade glioneuronal tumors not otherwise classified, we were able to refine the diagnosis for six patients. Finally, it is noteworthy that 30% of our Type I FCD cases (3 of 10 patients) showed evidence of a gain of the q-arm of chromosome 1. Partial chromosomal gains may be a previously underappreciated genetic mechanism of Type I FCD.

Altogether, genetic analysis of affected brain tissue is a valuable tool to understand the etiology of epilepsy-associated lesions and to refine diagnoses. It is conceivable that this information could be used in the future to inform patient care. For example, the presence of a particular variant at a certain allele fraction may inform on the expected course of disease and its severity, which could help providers determine the best treatment approaches. Likewise, knowledge of the underlying etiology could help to narrow down particular therapeutics most likely to succeed. In the future, genetic analysis of resected tissue may be considered standard of care. This study presents a framework for the detection of brain somatic variation in epilepsy-associated developmental lesions and proposes new genetic mechanisms of FCD for further study.

## Data Availability

All data produced in the present study are available upon reasonable request to the authors.

## Abbreviations

FCD: focal cortical dysplasia
LEAT: long-term epilepsy-associated tumor
ILAE: International League Against Epilepsy
VAF: variant allele fraction

## Acknowledgements

The authors are grateful to the patients and families who participated in this study.

## Supplementary material

Supplementary material is available online.

## References

1. Barba C, Specchio N, Guerrini R, et al. Increasing volume and complexity of pediatric epilepsy surgery with stable seizure outcome between 2008 and 2014: A nationwide multicenter study. Epilepsy Behav. 2017;75:151–157.

2. Blumcke I, Thom M, Aronica E, et al. The clinicopathologic spectrum of focal cortical dysplasias: a consensus classification proposed by an ad hoc Task Force of the ILAE Diagnostic Methods Commission. Epilepsia. 2011;52(1):158–174.

3. Blumcke I, Aronica E, Becker A, et al. Low-grade epilepsy-associated neuroepithelial tumours - the 2016 WHO classification. Nat Rev Neurol. 2016;12(12):732–740.

4. Slegers RJ, Blumcke I. Low-grade developmental and epilepsy associated brain tumors: a critical update 2020. Acta Neuropathol Commun. 2020;8(1):27.

5. Baldassari S, Ribierre T, Marsan E, et al. Dissecting the genetic basis of focal cortical dysplasia: a large cohort study. Acta Neuropathol. 2019;138(6):885–900.

6. Winawer MR, Griffin NG, Samanamud J, et al. Somatic SLC35A2 variants in the brain are associated with intractable neocortical epilepsy. Ann Neurol. 2018;83(6):1133–1146.

7. Sim NS, Seo Y, Lim JS, et al. Brain somatic mutations in SLC35A2 cause intractable epilepsy with aberrant N-glycosylation. Neurol Genet. 2018;4(6):e294.

8. Lim JS, Kim WI, Kang HC, et al. Brain somatic mutations in MTOR cause focal cortical dysplasia type II leading to intractable epilepsy. Nat Med. 2015;21(4):395–400.

9. Koh HY, Kim SH, Jang J, et al. BRAF somatic mutation contributes to intrinsic epileptogenicity in pediatric brain tumors. Nat Med. 2018;24(11):1662–1668.

10. Rivera B, Gayden T, Carrot-Zhang J, et al. Germline and somatic FGFR1 abnormalities in dysembryoplastic neuroepithelial tumors. Acta Neuropathol. 2016;131(6):847–863.

11. Kelly BJ, Fitch JR, Hu Y, et al. Churchill: an ultra-fast, deterministic, highly scalable and balanced parallelization strategy for the discovery of human genetic variation in clinical and population-scale genomics. Genome Biol. 2015;16:6.

12. DePristo MA, Banks E, Poplin R, et al. A framework for variation discovery and genotyping using next-generation DNA sequencing data. Nat Genet. 2011;43(5):491–498.

13. Cibulskis K, Lawrence MS, Carter SL, et al. Sensitive detection of somatic point mutations in impure and heterogeneous cancer samples. Nat Biotechnol. 2013;31(3):213–219.

14. Richards S, Aziz N, Bale S, et al. Standards and guidelines for the interpretation of sequence variants: a joint consensus recommendation of the American College of Medical Genetics and Genomics and the Association for Molecular Pathology. Genet Med. 2015;17(5):405–424.

15. Koboldt DC, Zhang Q, Larson DE, et al. VarScan 2: somatic mutation and copy number alteration discovery in cancer by exome sequencing. Genome Res. 2012;22(3):568–576.

16. Miller KE, Koboldt DC, Schieffer KM, et al. Somatic SLC35A2 mosaicism correlates with clinical findings in epilepsy brain tissue. Neurol Genet. 2020;6(4):e460.

17. Koboldt DC, Miller KE, Miller AR, et al. PTEN somatic mutations contribute to spectrum of cerebral overgrowth. Brain. 2021.

18. Dobin A, Davis CA, Schlesinger F, et al. STAR: ultrafast universal RNA-seq aligner. Bioinformatics. 2013;29(1):15–21.

19. LaHaye S, Fitch J, Voytovich K, et al. Discovery of clinically relevant fusions in pediatric cancer. BMC Genomics. 2021;In Press.

20. Tian L, Li Y, Edmonson MN, et al. CICERO: a versatile method for detecting complex and diverse driver fusions using cancer RNA sequencing data. Genome Biol. 2020;21(1):126.

21. Louis DN, Perry A, Wesseling P, et al. The 2021 WHO Classification of Tumors of the Central Nervous System: a summary. Neuro Oncol. 2021;23(8):1231–1251.

22. Louis DN, Aldape K, Brat DJ, et al. Announcing cIMPACT-NOW: the Consortium to Inform Molecular and Practical Approaches to CNS Tumor Taxonomy. Acta Neuropathol. 2017;133(1):1–3.

23. Kopanos C, Tsiolkas V, Kouris A, et al. VarSome: the human genomic variant search engine. Bioinformatics. 2019;35(11):1978–1980.

24. Kenney JW, Sorokina O, Genheden M, Sorokin A, Armstrong JD, Proud CG. Dynamics of elongation factor 2 kinase regulation in cortical neurons in response to synaptic activity. J Neurosci. 2015;35(7):3034–3047.

25. Heise C, Taha E, Murru L, et al. eEF2K/eEF2 Pathway Controls the Excitation/Inhibition Balance and Susceptibility to Epileptic Seizures. Cereb Cortex. 2017;27(3):2226–2248.

26. Karczewski KJ, Francioli LC, Tiao G, et al. The mutational constraint spectrum quantified from variation in 141,456 humans. Nature. 2020;581(7809):434–443.

27. Maes T, Barcelo A, Buesa C. Neuron navigator: a human gene family with homology to unc-53, a cell guidance gene from Caenorhabditis elegans. Genomics. 2002;80(1):21–30.

28. McNeill EM, Klockner-Bormann M, Roesler EC, Talton LE, Moechars D, Clagett-Dame M. Nav2 hypomorphic mutant mice are ataxic and exhibit abnormalities in cerebellar development. Dev Biol. 2011;353(2):331–343.

29. Fujita A, Higashijima T, Shirozu H, et al. Pathogenic variants of DYNC2H1, KIAA0556, and PTPN11 associated with hypothalamic hamartoma. Neurology. 2019;93(3):e237–e251.

30. Siegfried A, Cances C, Denuelle M, et al. Noonan syndrome, PTPN11 mutations, and brain tumors. A clinical report and review of the literature. Am J Med Genet A. 2017;173(4):1061–1065.

31. Zhu Y, Shen J, Sun T, et al. Loss of Shp2 within radial glia is associated with cerebral cortical dysplasia, glial defects of cerebellum and impaired sensorymotor development in newborn mice. Mol Med Rep. 2018;17(2):3170–3177.

32. Fukata Y, Adesnik H, Iwanaga T, Bredt DS, Nicoll RA, Fukata M. Epilepsy-related ligand/receptor complex LGI1 and ADAM22 regulate synaptic transmission. Science. 2006;313(5794):1792–1795.

33. Dibbens LM, de Vries B, Donatello S, et al. Mutations in DEPDC5 cause familial focal epilepsy with variable foci. Nat Genet. 2013;45(5):546–551.

34. Zagaglia S, Selch C, Nisevic JR, et al. Neurologic phenotypes associated with COL4A1/2 mutations: Expanding the spectrum of disease. Neurology. 2018;91(22):e2078–e2088.

35. Ricos MG, Hodgson BL, Pippucci T, et al. Mutations in the mammalian target of rapamycin pathway regulators NPRL2 and NPRL3 cause focal epilepsy. Ann Neurol. 2016;79(1):120–131.

36. Terrone G, Marchese F, Vari MS, et al. A further contribution to the delineation of epileptic phenotype in PACS2-related syndrome. Seizure. 2020;79:53–55.

37. Tajan M, de Rocca Serra A, Valet P, Edouard T, Yart A. SHP2 sails from physiology to pathology. Eur J Med Genet. 2015;58(10):509–525.

