## Supplemental Table 1 for "Detection of brain somatic variation in epilepsy-associated developmental lesions"

| **Patient ID** | **Sex** | **MRI** | **Intracranial EEG** | **Surgery** | **Engel Class** | **Diagnosis** | **Neuropathology** | **Somatic Finding** | **Germline Finding** |
| --- | --- | --- | --- | --- | --- | --- | --- | --- | --- |
| 4 | M | Lesional |  | Right temporal lobectomy | IA | LEAT | Diffuse low-grade glioma (MAPK pathway-altered) | *FGFR1-TACC1* |  |
| 50 | F | Lesional | ECOG | Left temporal lesionectomy | IA | LEAT | Diffuse low-grade glioma (MAPK pathway-altered) and FCDIIIb | BRAF p. V600E |  |
| 2 | M | Lesional | ECOG | Right anterior temporal lobectomy with amygdalohippocampectomy | IA | LEAT | DNET | FGFR1 ITD |  |
| 5 | M | Lesional | ECOG | Left temporal lesionectomy | IA | LEAT | DNET | FGFR1 ITD |  |
| 9 | M | Lesional | ECOG | Left parietal lesionectomy | IA | LEAT | DNET | FGFR1 ITD |  |
| 28 | M | Lesional | ECOG | Left temporal lesionectomy | IIA | LEAT | DNET | FGFR1 ITD / ADAM22 |  |
| 7 | M | Lesional | ECOG | Right temporal lesionectomy | IA | LEAT | DNET and FCDIIIb | *KRAS* |  |
| 41 | M | Lesional | ECOG | Left frontal lesionectomy | IA | LEAT | DNET and FCDIIIb | FGFR1 ITD |  |
| 1 | F | Lesional |  | Right parietal lesionectomy | IA | LEAT | Ganglioglioma | BRAF p. V600E |  |
| 6 | M | Lesional | ECOG | Right anterior temporal lobectomy with amygdalohippocampectomy | IA | LEAT | Ganglioglioma | BRAF p. V600E |  |
| 45 | M | Lesional | ECOG | Right temporal lesionectomy | IA | LEAT | Ganglioglioma | BRAF p. V600E |  |
| 3 | M | Lesional | Grid | Right temporo-occipital lobectomy and inferior parietal lobule corticectomy | IA | LEAT | PLNTY and FCDIIIb | *FGFR2-CLIP1* |  |
| 34 | M | Lesional | Grid | Left anterior temporal lobectomy with amygdalohippocampectomy | IA | LEAT | PLNTY and FCDIIIb | *FGFR1-INA* |  |
| 46 | F | Lesional | SEEG | Left temporal lesionectomy | IA | LEAT | PLNTY and FCDIIIb | *FGFR2-PASD1* |  |
| 8 | M | Lesional |  | Right temporo-parieto-occipital lesionectomy | IA | LEAT | PXA | BRAF p. V600E |  |
| 10 | F | Lesional |  | Right frontal lesionectomy | IA | LEAT | PXA | BRAF p. V600E |  |
| 26 | F | Lesional | SEEG | Left parieto-occipital lobectomy | IA | MCD | FCDIa |  |  |
| 36 | F | Non-lesional | SEEG | Right frontal lesionectomy | IA | MCD | FCDIa |  |  |
| 40 | M | Non-lesional | SEEG | Right frontal lesionectomy | IA | MCD | FCDIa |  | *NPRL3* |
| 16 | F | Lesional |  | Right functional hemispherectomy | IA | MCD | FCDIb |  |  |
| 11 | M | Lesional | Grid | Left temporo-occipital lobectomy and inferior parietal lobule corticectomy | IA | MCD | FCDIc | *SLC35A2* |  |
| 15 | F | Lesional | ECOG | Right anterior temporal lobectomy | IC | MCD | FCDIc |  |  |
| 20 | F | Lesional | Grid | Right anatomic hemispherectomy | IIA | MCD | FCDIc | *EEF2* |  |
| 24 | F | Lesional |  | Right functional hemispherectomy | IA | MCD | FCDIc | Chr 1q |  |
| 35 | F | Lesional | ECOG | Right functional hemispherectomy | IA | MCD | FCDIc | Chr 1q |  |
| 39 | M | Lesional | Grid | Right temporal lobectomy and inferior parietal lobule corticectomy | IIIA | MCD | FCDIc |  | *PACS2* |
| 43 | M | Lesional |  | Left functional hemispherotomy | IIIA | MCD | FCDIc | Chr 1q |  |
| 44 | M | Lesional |  | Right functional hemispherectomy | IA | MCD | FCDIc |  | *DEPDC5* |
| 48 | M | Non-lesional | SEEG | Right frontal lesionectomy | IIB | MCD | FCDIc | *SLC35A2* |  |
| 13 | F | Non-lesional | Grid | Left temporo-occipital lesionectomy | IB | MCD | FCDIIa |  |  |
| 17 | M | Lesional |  | Left anatomic hemispherectomy | IA | MCD | FCDIIa | *PTEN* |  |
| 21 | F | Lesional |  | Right functional hemispherectomy | ID | MCD | FCDIIa |  |  |
| 33 | F | Non-lesional | SEEG | Right frontal lesionecomty | IA | MCD | FCDIIa |  |  |
| 47 | M | Lesional | ECOG | Left functional hemispherotomy | IID | MCD | FCDIIa | *RHEB* |  |
| 25 | M | Lesional | ECOG | Left frontal lesionectomy | IA | MCD | FCDIIb |  |  |
| 27 | F | Lesional | ECOG | Left parietal lesionectomy | IA | MCD | FCDIIb |  |  |
| 29 | M | Lesional | ECOG | Left frontal lesionectomy | IA | MCD | FCDIIb | *MTOR* |  |
| 42 | F | Non-lesional | SEEG | Right temporal lesionectomy | IA | MCD | FCDIIb |  |  |
| 19 | F | Lesional | ECOG | Left anterior temporal lobectomy with amygdalohippocampectomy | IA | MCD | FCDIIIa |  |  |
| 38 | F | Lesional |  | Left functional hemispherotomy | IA | MCD | FCDIIIa | *PTPN11* |  |
| 32 | M | Lesional | ECOG | Right temporal lesionectomy | IA | MCD | FCDIIIc |  |  |
| 12 | M | Lesional |  | Left anatomic hemispherectomy | IA | MCD | FCDIIId |  |  |
| 14 | M | Lesional | SEEG | Left anterior temporal lobectomy with amygdalohippocampectomy | IVA | MCD | FCDIIId |  |  |
| 18 | F | Lesional |  | Right anatomic hemispherectomy | IVA | MCD | FCDIIId |  |  |
| 22 | F | Lesional |  | Right functional hemispherectomy | IA | MCD | FCDIIId |  |  |
| 23 | M | Lesional |  | Left functional hemispherotomy | IA | MCD | FCDIIId | *NAV2* |  |
| 30 | F | Lesional |  | Right functional hemispherectomy | IA | MCD | FCDIIId |  |  |
| 31 | F | Lesional |  | Left functional hemispherotomy | IA | MCD | FCDIIId |  |  |
| 37 | F | Lesional |  | Left functional hemispherotomy | IA | MCD | FCDIIId |  | *COL4A1* |
| 49 | F | Lesional |  | Left functional hemispherotomy | IA | MCD | FCDIIId | *PTPN11* |  |
