## Supplemental Table 2 for "Detection of brain somatic variation in epilepsy-associated developmental lesions"

| **Exome Sequencing - Somatic Findings** | | | | | | | |
| --- | --- | --- | --- | --- | --- | --- | --- |
| **Long-term Epilepsy-Associated Tumors** | | | | | | | |
| **Patient ID** | **Neuropathology** | **Gene** | **Variant Description** | **Tissue Samples Sequenced** | **Observed in tumor (VAF %)** | **Observed in adjacent dysplasia (VAF %)** | **Observed in mixed tumor/dysplasia (VAF %)** |
| 5 | DNET | FGFR1 ITD | exon 9 - exon 18 | 1 | NA | NA | Yes |
| 9 | DNET | FGFR1 ITD | exon 9 - exon 18 | 1 | Yes | NA | NA |
| 41 | DNET and FCDIIIb | FGFR1 ITD | exon 10 - exon 18 | 3 | Yes | No (0.0) | NA |
| 28 | DNET | FGFR1 ITD | exon 10 - exon 18 | 1 | Yes | NA | NA |
| 28 | DNET | *ADAM22* | NM_021721.4:c.42_43delCT:p.Val16Profs*71 | 1 | Yes (17.9) | NA | NA |
| 2 | DNET | FGFR1 ITD | exon 10 - exon 18 | 1 | Yes | NA | NA |
| 7 | DNET and FCDIIIb | *KRAS* | NM_004985.5:c.181C>A:p.Gln61Lys | 5 | Yes (0.4 - 49.2) | NA | No (0.0) |
| 1 | Ganglioglioma | *BRAF* | NM_004333.6:c.1799T>A:p.Val600Glu | 1 | Yes (17.4) | NA | NA |
| 6 | Ganglioglioma | *BRAF* | NM_004333.6:c.1799T>A:p.Val600Glu | 1 | Yes (28.3) | NA | NA |
| 45 | Ganglioglioma | *BRAF* | NM_004333.6:c.1799T>A:p.Val600Glu | 1 | Yes (32.0) | NA | NA |
| 50 | Diffuse low-grade glioma (MAPK pathway-altered) and FCDIIIb | *BRAF* | NM_004333.6:c.1799T>A:p.Val600Glu | 2 | NA | NA | Yes (0.0 - 12.7) |
| 4 | Diffuse low-grade glioma (MAPK pathway-altered) | *FGFR1-TACC1* | FGFR1 (8:38413918-) + TACC1 (8:38836162+) | 1 | Yes | NA | NA |
| 34 | PLNTY and FCDIIIb | *FGFR2-INA* | FGFR2 (10:121483698-) + INA (10:103287035+) | 9 | Yes | No (0.0) | Yes |
| 3 | PLNTY and FCDIIIb | *FGFR2-CLIP1* | FGFR2 (10:121483698-) + CLIP1 (12:122288541-) | 8 | Yes | No (0.0) | NA |
| 46 | PLNTY and FCDIIIb | *FGFR2-PASD1* | FGFR2 (10:121483698-) + PASD1 (X:151659713+) | 3 | Yes | No (0.0) | NA |
| 8 | PXA | *BRAF* | NM_004333.6:c.1799T>A:p.Val600Glu | 1 | Yes (34.5) | NA | NA |
| 10 | PXA | *BRAF* | NM_004333.6:c.1799T>A:p.Val600Glu | 1 | Yes (40.5) | NA | NA |
| **Focal Cortical Dysplasia** | | | | | | | |
| **Patient ID** | **Neuropathology** | **Gene** | **Variant Description** | **Tissue Samples Sequenced** | **VAF %** |  |  |
| 24 | FCDIc | Chr1 | Chr1q Gain | 6 | 0.1 - 13.7 |  |  |
| 43 | FCDIc | Chr1 | Chr1q Gain | 5 | 0.4 - 20.2 |  |  |
| 35 | FCDIc | Chr1 | Chr1q Gain | 1 | 20.0 - 30.0 |  |  |
| 20 | FCDIc | *EEF2* | NM_001961.4:c.1132G>A:p.Asp378Asn | 4 | 0.3 - 4.0 |  |  |
| 11 | FCDIc | *SLC35A2* | NM_005660.3:c.634_635del:p.Ser212Leufs*9 | 13 | 0.6 - 27.7 |  |  |
| 48 | FCDIc | *SLC35A2* | NM_005660.3:c.168C>G:p.Tyr56* | 4 | 0 - 8.8 |  |  |
| 17 | FCDIIa | *PTEN* | NM_000314.8:c.255_262delinsC:p.Ala86Ilefs*11 | 1 | 33.3 |  |  |
| 17 | FCDIIa | *PTEN* | NM_000314.8:c.1110_1111dup:p.Asp371Valfs*46 | 1 | 10.6 |  |  |
| 47 | FCDIIa | *RHEB* | NM_005614.4:c.104_105delinsTG:p.Tyr35Leu | 4 | 5.9 - 11.0 |  |  |
| 29 | FCDIIb | *MTOR* | NM_004958.4:c.6644C>A:p.Ser2215Tyr | 1 | 1.9 |  |  |
| 38 | FCDIIIa | *PTPN11* | NM_002834.5:c.1502G>A:p.Arg501Lys | 2 | 0 - 3.0 |  |  |
| 23 | FCDIIId | *NAV2* | NM_182964.6:c.1093G>A:p.Val365Met | 4 | 0 - 7.9 |  |  |
| 49 | FCDIIId | *PTPN11* | NM_002834.5:c.50A>G:p.Glu17Gly | 3 | 0 - 7.3 |  |  |
| **Exome Sequencing - Germline Findings** | | | | | | | |
| **Focal Cortical Dysplasia** | | | | | | | |
| **Patient ID** | **Neuropathology** | **Gene** | **Variant Description** | **Tissue Samples Sequenced** | **Blood VAF** | **ACMG/AMP Criteria** |  |
| 40 | FCDIa | *NPRL3* | NM_001077350.3:c.995_997delinsCA:p.Asn332Thrfs*81 | 1 | 32% | LP (PVS1, PM2) |  |
| 44 | FCDIc | *DEPDC5* | NM_001242896.3:c.346C>T:p.Arg116* | 3 | 41% | P (PVS1, PP3, PP5) |  |
| 39 | FCDIc | *PACS2* | NM_001100913.3:c.2147del:p.Gly716Alafs*29 | 5 | 49% | VUS (PM2) |  |
| 37 | FCDIIId | *COL4A1* | NM_001845.6:c.1258G>A:p.Gly420Arg | 10 | 43% | LP (PM1, PM2, PP2, PP3) |  |
