## Supplemental Table 3 for "Detection of brain somatic variation in epilepsy-associated developmental lesions"

| **Patient ID** | **Gene** | **HGVS** | **Tissue Origin** | **VAF_WES** | **VAF_targeted** | **Targeted_result** |
| --- | --- | --- | --- | --- | --- | --- |
| 11 | SLC35A2 | NM_005660.3:c.634_635del:p.Ser212Leufs*9 | Blood | 0.0% | 0.2% | refuted |
| 11 | SLC35A2 | NM_005660.3:c.634_635del:p.Ser212Leufs*9 | Left superior temporal gyrus | NA | 5.3% | validated |
| 11 | SLC35A2 | NM_005660.3:c.634_635del:p.Ser212Leufs*9 | Left inferior occipital lobe | NA | 1.7% | validated |
| 11 | SLC35A2 | NM_005660.3:c.634_635del:p.Ser212Leufs*9 | Left medial occipital lobe | NA | 10.3% | validated |
| 11 | SLC35A2 | NM_005660.3:c.634_635del:p.Ser212Leufs*9 | Left medial occipital lobe | NA | 11.8% | validated |
| 11 | SLC35A2 | NM_005660.3:c.634_635del:p.Ser212Leufs*9 | Left inferior parietal cortex | NA | 8.8% | validated |
| 11 | SLC35A2 | NM_005660.3:c.634_635del:p.Ser212Leufs*9 | Left inferior parietal cortex | NA | 8.7% | validated |
| 11 | SLC35A2 | NM_005660.3:c.634_635del:p.Ser212Leufs*9 | Left temporal lobe | 3.0% | 1.9% | validated |
| 11 | SLC35A2 | NM_005660.3:c.634_635del:p.Ser212Leufs*9 | Left temporal lobe | 3.3% | 2.7% | validated |
| 11 | SLC35A2 | NM_005660.3:c.634_635del:p.Ser212Leufs*9 | Left amygdala | 12.5% | 5.2% | validated |
| 11 | SLC35A2 | NM_005660.3:c.634_635del:p.Ser212Leufs*9 | Left hippocampus | 27.7% | 13.5% | validated |
| 11 | SLC35A2 | NM_005660.3:c.634_635del:p.Ser212Leufs*9 | Left occipital lobe and pole | 9.8% | 2.8% | validated |
| 11 | SLC35A2 | NM_005660.3:c.634_635del:p.Ser212Leufs*9 | Left occipital lobe and pole | 0.6% | 2.3% | validated |
| 11 | VEGFC | NM_005429.4:c.1220G>A:p.Arg407His | Left inferior occipital lobe | NA | 0.4% | refuted |
| 11 | VEGFC | NM_005429.4:c.1220G>A:p.Arg407His | Left medial occipital lobe | NA | 0.1% | refuted |
| 11 | VEGFC | NM_005429.4:c.1220G>A:p.Arg407His | Left medial occipital lobe | NA | 0.3% | refuted |
| 11 | VEGFC | NM_005429.4:c.1220G>A:p.Arg407His | Left inferior parietal cortex | NA | 0.3% | refuted |
| 11 | VEGFC | NM_005429.4:c.1220G>A:p.Arg407His | Left inferior parietal cortex | NA | 0.2% | refuted |
| 11 | VEGFC | NM_005429.4:c.1220G>A:p.Arg407His | Left superior temporal gyrus | NA | 0.4% | refuted |
| 11 | VEGFC | NM_005429.4:c.1220G>A:p.Arg407His | Blood | 0.00% | 0.1% | refuted |
| 11 | VEGFC | NM_005429.4:c.1220G>A:p.Arg407His | Left temporal lobe | 0.46% | 0.3% | refuted |
| 11 | VEGFC | NM_005429.4:c.1220G>A:p.Arg407His | Left temporal lobe | 0.00% | 0.2% | refuted |
| 11 | VEGFC | NM_005429.4:c.1220G>A:p.Arg407His | Left amygdala | 2.40% | 0.4% | refuted |
| 11 | VEGFC | NM_005429.4:c.1220G>A:p.Arg407His | Left hippocampus | 2.30% | 0.5% | refuted |
| 11 | VEGFC | NM_005429.4:c.1220G>A:p.Arg407His | Left occipital lobe and pole | 0.87% | 0.4% | refuted |
| 11 | VEGFC | NM_005429.4:c.1220G>A:p.Arg407His | Left occipital lobe and pole | 0.00% | 0.4% | refuted |
| 13 | SDHA | NM_001294332.1:c.1408-3C>A | Left posterior-inferior temporal gyrus and occipitotemporal gyrus | 1.84% | 0.0% | refuted |
| 13 | SDHA | NM_001294332.1:c.1408-3C>A | Left posterior-inferior temporal gyrus and occipitotemporal gyrus | 0.00% | 0.0% | refuted |
| 17 | PTEN | NM_000314.8:c.1110_1111dup:p.Asp371Valfs*46 | Left temporal pole | NA | 0.0% | refuted |
| 17 | PTEN | NM_000314.8:c.1110_1111dup:p.Asp371Valfs*46 | Left frontal lobe | NA | 0.0% | refuted |
| 17 | PTEN | NM_000314.8:c.1110_1111dup:p.Asp371Valfs*46 | Left frontal pole | NA | 0.0% | refuted |
| 17 | PTEN | NM_000314.8:c.1110_1111dup:p.Asp371Valfs*46 | Left frontal lobe | NA | 0.0% | refuted |
| 17 | PTEN | NM_000314.8:c.1110_1111dup:p.Asp371Valfs*46 | Left anterior frontal lobe | NA | 0.0% | refuted |
| 17 | PTEN | NM_000314.8:c.1110_1111dup:p.Asp371Valfs*46 | Blood | 0.00% | 0.0% | refuted |
| 17 | PTEN | NM_000314.8:c.1110_1111dup:p.Asp371Valfs*46 | Left lateral posterior temporal lobe | NA | 19.2% | validated |
| 17 | PTEN | NM_000314.8:c.1110_1111dup:p.Asp371Valfs*46 | Left lateral anterior temporal lobe | NA | 21.4% | validated |
| 17 | PTEN | NM_000314.8:c.1110_1111dup:p.Asp371Valfs*46 | Left posterior lateral temporal lobe | NA | 13.3% | validated |
| 17 | PTEN | NM_000314.8:c.1110_1111dup:p.Asp371Valfs*46 | Left parietal lobe | NA | 0.5% | validated |
| 17 | PTEN | NM_000314.8:c.1110_1111dup:p.Asp371Valfs*46 | Left medial temporal lobe | NA | 0.9% | validated |
| 17 | PTEN | NM_000314.8:c.1110_1111dup:p.Asp371Valfs*46 | Left parietal lobe | NA | 17.0% | validated |
| 17 | PTEN | NM_000314.8:c.1110_1111dup:p.Asp371Valfs*46 | Left posterior parietal lobe | NA | 23.0% | validated |
| 17 | PTEN | NM_000314.8:c.1110_1111dup:p.Asp371Valfs*46 | Left posterior parietal lobe | NA | 26.1% | validated |
| 17 | PTEN | NM_000314.8:c.1110_1111dup:p.Asp371Valfs*46 | Left occipital | NA | 20.0% | validated |
| 17 | PTEN | NM_000314.8:c.255_262delinsC:p.Ala86Ilefs*11 | Left temporal pole | NA | 19.3% | validated |
| 17 | PTEN | NM_000314.8:c.255_262delinsC:p.Ala86Ilefs*11 | Left frontal lobe | NA | 21.4% | validated |
| 17 | PTEN | NM_000314.8:c.255_262delinsC:p.Ala86Ilefs*11 | Left lateral anterior temporal lobe | NA | 21.1% | validated |
| 17 | PTEN | NM_000314.8:c.255_262delinsC:p.Ala86Ilefs*11 | Left frontal pole | NA | 16.6% | validated |
| 17 | PTEN | NM_000314.8:c.255_262delinsC:p.Ala86Ilefs*11 | Left frontal lobe | NA | 14.5% | validated |
| 17 | PTEN | NM_000314.8:c.255_262delinsC:p.Ala86Ilefs*11 | Left posterior lateral temporal lobe | NA | 29.0% | validated |
| 17 | PTEN | NM_000314.8:c.255_262delinsC:p.Ala86Ilefs*11 | Left parietal lobe | NA | 15.3% | validated |
| 17 | PTEN | NM_000314.8:c.255_262delinsC:p.Ala86Ilefs*11 | Left medial temporal lobe | NA | 19.8% | validated |
| 17 | PTEN | NM_000314.8:c.255_262delinsC:p.Ala86Ilefs*11 | Left anterior frontal lobe | NA | 15.9% | validated |
| 17 | PTEN | NM_000314.8:c.255_262delinsC:p.Ala86Ilefs*11 | Left parietal lobe | NA | 23.3% | validated |
| 17 | PTEN | NM_000314.8:c.255_262delinsC:p.Ala86Ilefs*11 | Left posterior parietal lobe | NA | 25.7% | validated |
| 17 | PTEN | NM_000314.8:c.255_262delinsC:p.Ala86Ilefs*11 | Left posterior parietal lobe | NA | 26.8% | validated |
| 17 | PTEN | NM_000314.8:c.255_262delinsC:p.Ala86Ilefs*11 | Left occipital | NA | 31.1% | validated |
| 17 | PTEN | NM_000314.8:c.255_262delinsC:p.Ala86Ilefs*11 | Left lateral posterior temporal lobe | NA | 30.4% | validated |
| 17 | PTEN | NM_000314.8:c.1110_1111dup:p.Asp371Valfs*46 | Left frontal lobe | 10.6% | 13.6% | validated |
| 17 | PTEN | NM_000314.8:c.255_262delinsC:p.Ala86Ilefs*11 | Blood | 0.00% | 2.9% | validated |
| 17 | PTEN | NM_000314.8:c.255_262delinsC:p.Ala86Ilefs*11 | Left frontal lobe | 33.30% | 21.7% | validated |
| 18 | PTPN11 | NM_002834.3:c.1492C>T:p.Arg498Trp | Blood | 0.00% | 0.2% | refuted |
| 18 | PTPN11 | NM_002834.3:c.1492C>T:p.Arg498Trp | Hippocampus | 2.21% | 0.3% | refuted |
| 18 | PTPN11 | NM_002834.3:c.1492C>T:p.Arg498Trp | Right frontal lobe | 0.00% | 0.1% | refuted |
| 18 | PTPN11 | NM_002834.3:c.1492C>T:p.Arg498Trp | Medial frontal lobe | NA | 0.0% | refuted |
| 18 | PTPN11 | NM_002834.3:c.1492C>T:p.Arg498Trp | Posterior frontal lobe | NA | 0.1% | refuted |
| 18 | PTPN11 | NM_002834.3:c.1492C>T:p.Arg498Trp | Parietal lobe | NA | 0.2% | refuted |
| 20 | EEF2 | NM_001961.3:c.1132G>A:p.Asp378Asn | Blood | 0.00% | 0.0% | refuted |
| 20 | EEF2 | NM_001961.3:c.1132G>A:p.Asp378Asn | Right occipital lobe | 0.33% | 0.1% | validated |
| 20 | EEF2 | NM_001961.3:c.1132G>A:p.Asp378Asn | Right parietal lobe | 0.81% | 1.4% | validated |
| 20 | EEF2 | NM_001961.3:c.1132G>A:p.Asp378Asn | Right posterior frontal | 4.01% | 3.2% | validated |
| 20 | EEF2 | NM_001961.3:c.1132G>A:p.Asp378Asn | Right anterior frontal | 0.56% | 0.7% | validated |
| 22 | IGF1R | NM_000875.4:c.3846C>A:p.Ser1282Arg | Blood | 0.00% | 0.0% | refuted |
| 22 | IGF1R | NM_000875.4:c.3846C>A:p.Ser1282Arg | Right frontal operculum | 0.30% | 0.0% | refuted |
| 22 | IGF1R | NM_000875.4:c.3846C>A:p.Ser1282Arg | Right temporal operculum | 1.46% | 0.0% | refuted |
| 22 | IGF1R | NM_000875.4:c.3846C>A:p.Ser1282Arg | Right anterior temporal lobe; Lateral neocortex | 0.00% | 0.0% | refuted |
| 23 | NAV2 | NM_145117.4:c.1093G>A:p.Val365Met | Blood | 0.00% | 0.0% | refuted |
| 23 | NAV2 | NM_145117.4:c.1093G>A:p.Val365Met | Left anterior temporal lobe; Lateral cortex | 0.00% | 0.0% | refuted |
| 23 | NAV2 | NM_145117.4:c.1093G>A:p.Val365Met | Left pars orbitalis | 7.88% | 7.9% | validated |
| 23 | NAV2 | NM_145117.4:c.1093G>A:p.Val365Met | Left frontal operculum | 0.74% | 0.9% | validated |
| 23 | NAV2 | NM_145117.4:c.1093G>A:p.Val365Met | Left parietal operculum | 0.00% | 0.5% | validated |
| 24 | DYNC1H1 | NM_001376.4:c.9515C>T:p.Thr3172Ile | Blood | 0.00% | 0.1% | refuted |
| 24 | DYNC1H1 | NM_001376.4:c.9515C>T:p.Thr3172Ile | Right frontal operculum | 0.00% | 0.1% | refuted |
| 24 | DYNC1H1 | NM_001376.4:c.9515C>T:p.Thr3172Ile | Right inferior posterior frontal gyrus | 5.13% | 0.1% | refuted |
| 24 | DYNC1H1 | NM_001376.4:c.9515C>T:p.Thr3172Ile | Right temporal lobe; Lateral cortex | 0.00% | 0.1% | refuted |
| 24 | DYNC1H1 | NM_001376.4:c.9515C>T:p.Thr3172Ile | Right amygdala | 0.50% | 0.1% | refuted |
| 24 | DYNC1H1 | NM_001376.4:c.9515C>T:p.Thr3172Ile | Right hippocampus | 0.28% | 0.1% | refuted |
| 28 | ADAM22 | NM_021721.4:c.42_43del:p.Val16Profs*71 | Blood | 0.00% | 0.0% | refuted |
| 28 | ADAM22 | NM_021721.4:c.42_43del:p.Val16Profs*71 | Left temporal | 17.89% | 15.6% | validated |
| 28 | APC2 | NM_005883.2:c.3636G>T:p.Lys1212Asn | Blood | 0.00% | 0.0% | refuted |
| 28 | APC2 | NM_005883.2:c.3636G>T:p.Lys1212Asn | Left temporal | 3.33% | 0.0% | refuted |
| 28 | CCDC88A | NM_018084.4:c.1223T>A:p.Leu408* | Blood | 0.00% | 0.0% | refuted |
| 28 | CCDC88A | NM_018084.4:c.1223T>A:p.Leu408* | Left temporal | 1.99% | 0.1% | refuted |
| 28 | GABRB3 | NM_000814.5:c.1393T>A:p.Leu465Ile | Blood | 0.00% | 0.0% | refuted |
| 28 | GABRB3 | NM_000814.5:c.1393T>A:p.Leu465Ile | Left temporal | 2.30% | 0.0% | refuted |
| 28 | RYR2 | NM_001035.2:c.12283G>T:p.Gly4095Cys | Blood | 1.06% | 0.0% | refuted |
| 28 | RYR2 | NM_001035.2:c.12283G>T:p.Gly4095Cys | Left temporal | 5.26% | 0.0% | refuted |
| 28 | TARDBP | NM_007375.3:c.1004G>A:p.Gly335Asp | Blood | 0.00% | 0.0% | refuted |
| 28 | TARDBP | NM_007375.3:c.1004G>A:p.Gly335Asp | Left temporal | 2.08% | 0.0% | refuted |
| 29 | MTOR | NM_004958.3:c.6644C>A:p.Ser2215Tyr | Blood | 0.00% | 0.0% | refuted |
| 29 | MTOR | NM_004958.3:c.6644C>A:p.Ser2215Tyr | Left inferior precentral gyrus and postcentral gyrus | 1.88% | 1.6% | validated |
| 30 | KIF22 | NM_007317.3:c.67_68del:p.Ser23Argfs*39 | Blood | 0.00% | NA | NA |
| 30 | KIF22 | NM_007317.3:c.67_68del:p.Ser23Argfs*39 | Right temporal lobe; Lateral neocortex | 1.19% | 0.0% | refuted |
| 30 | KIF22 | NM_007317.3:c.67_68del:p.Ser23Argfs*39 | Right hippocampus | 0.90% | 0.0% | refuted |
| 34 | KMT2C | NM_170606.3:c.6577_6578delinsGG:p.Pro2193Gly | Blood | 0.00% | NA | NA |
| 34 | KMT2C | NM_170606.3:c.6577_6578delinsGG:p.Pro2193Gly | Left temporal lobe | 0.60% | NA | NA |
| 34 | KMT2C | NM_170606.3:c.6577_6578delinsGG:p.Pro2193Gly | Left temporal lobe | 0.28% | NA | NA |
| 34 | KMT2C | NM_170606.3:c.6577_6578delinsGG:p.Pro2193Gly | Superior temporal lobe | 0.76% | NA | NA |
| 34 | KMT2C | NM_170606.3:c.6577_6578delinsGG:p.Pro2193Gly | Amygdala | 0.27% | NA | NA |
| 34 | KMT2C | NM_170606.3:c.6577_6578delinsGG:p.Pro2193Gly | Posterior left temporal lobe | 0.83% | NA | NA |
| 34 | KMT2C | NM_170606.3:c.6577_6578delinsGG:p.Pro2193Gly | Hippocampus | 0.00% | NA | NA |
| 34 | KMT2C | NM_170606.3:c.6577_6578delinsGG:p.Pro2193Gly | Anterior temporal | 0.65% | 0.0% | refuted |
| 34 | KMT2C | NM_170606.3:c.6577_6578delinsGG:p.Pro2193Gly | Left temporal lobe | 0.26% | 0.0% | refuted |
| 34 | KMT2C | NM_170606.3:c.6577_6578delinsGG:p.Pro2193Gly | Left anterior temporal lobe | 0.42% | 0.0% | refuted |
| 36 | POLR1A | NM_015425.4:c.4832C>T:p.Thr1611Met | Blood | 0.00% | NA | NA |
| 36 | POLR1A | NM_015425.4:c.4832C>T:p.Thr1611Met | Right middle frontal gyrus | 2.68% | 2.7% | validated |
| 36 | POLR1A | NM_015425.4:c.4832C>T:p.Thr1611Met | Anterior precentral gyrus | 0.75% | 2.1% | validated |
| 36 | POLR1A | NM_015425.4:c.4832C>T:p.Thr1611Met | Superior frontal gyrus | 1.31% | 3.0% | validated |
| 38 | PTPN11 | NM_002834.5:c.1502G>A:p.Arg501Lys | Blood | 0.00% | 0.0% | refuted |
| 38 | PTPN11 | NM_002834.5:c.1502G>A:p.Arg501Lys | Left inferior parietal region | 0.00% | 0.0% | refuted |
| 38 | PTPN11 | NM_002834.5:c.1502G>A:p.Arg501Lys | Left superior temporal gyrus | NA | 0.0% | refuted |
| 38 | PTPN11 | NM_002834.5:c.1502G>A:p.Arg501Lys | Left posterior inferior frontal region | 2.96% | 3.3% | validated |
| 41 | SLC26A1 | NM_022042.3:c.1982del:p.Gly661Alafs*66 | Blood | 0.00% | 0.0% | refuted |
| 41 | SLC26A1 | NM_022042.3:c.1982del:p.Gly661Alafs*66 | Left frontal tumor | 0.00% | 0.0% | refuted |
| 41 | SLC26A1 | NM_022042.3:c.1982del:p.Gly661Alafs*66 | Middle frontal tumor | 0.00% | 0.0% | refuted |
| 41 | SLC26A1 | NM_022042.3:c.1982del:p.Gly661Alafs*66 | Lateral superior frontal | 4.17% | 3.1% | validated |
| 42 | RAI1 | NM_030665.3:c.2204C>A:p.Ala735Asp | Blood | 0.00% | 0.0% | refuted |
| 42 | RAI1 | NM_030665.3:c.2204C>A:p.Ala735Asp | Right anterior temporal lobe | 0.90% | 0.0% | refuted |
| 42 | RAI1 | NM_030665.3:c.2204C>A:p.Ala735Asp | Right posterior temporal lobe | 1.42% | 0.0% | refuted |
| 44 | ENPP1 | NM_006208.3:c.2200A>T:p.Lys734* | Blood | 0.00% | 0.6% | validated |
| 44 | ENPP1 | NM_006208.3:c.2200A>T:p.Lys734* | Right frontal operculum | 2.98% | 3.3% | validated |
| 44 | ENPP1 | NM_006208.3:c.2200A>T:p.Lys734* | Right anterior frontal lobe | 2.12% | 1.5% | validated |
| 44 | ENPP1 | NM_006208.3:c.2200A>T:p.Lys734* | Right superior frontal lobe | 5.30% | 3.3% | validated |
| 45 | PTPN11 | NM_002834.5:c.1492C>T:p.Arg498Trp | Blood | 0.00% | 0.7% | refuted |
| 45 | PTPN11 | NM_002834.5:c.1492C>T:p.Arg498Trp | Right temporal | 2.75% | 0.5% | refuted |
| 45 | BRAF | NM_004333.6:c.1799T>A:p.Val600Glu | Blood | 0.00% | 0.0% | refuted |
| 45 | BRAF | NM_004333.6:c.1799T>A:p.Val600Glu | Tumor | 32.01% | 36.3% | validated |
| 47 | RHEB | NM_005614.4:c.104_105delinsTG:p.Tyr35Leu | Inferior temporal parietal | 5.15% | NA | NA |
| 47 | RHEB | NM_005614.4:c.104_105delinsTG:p.Tyr35Leu | Blood | 0.00% | 0.0% | refuted |
| 47 | RHEB | NM_005614.4:c.104_105delinsTG:p.Tyr35Leu | Temporal lobe | 11.40% | 12.2% | validated |
| 47 | RHEB | NM_005614.4:c.104_105delinsTG:p.Tyr35Leu | Unspecified | 9.71% | 13.3% | validated |
| 47 | RHEB | NM_005614.4:c.104_105delinsTG:p.Tyr35Leu | Anterior parietal temporal | 6.59% | 6.2% | validated |
| 48 | SLC35A2 | NM_005660.3:c.168C>G:p.Tyr56* | Blood | 0.00% | 0.0% | refuted |
| 48 | SLC35A2 | NM_005660.3:c.168C>G:p.Tyr56* | Anterior resection boundary | 0.00% | 0.1% | refuted |
| 48 | SLC35A2 | NM_005660.3:c.168C>G:p.Tyr56* | Anterior resection boundary | 0.00% | 0.0% | refuted |
| 48 | SLC35A2 | NM_005660.3:c.168C>G:p.Tyr56* | Anterior superior frontal gyrus | 8.76% | 12.4% | validated |
| 48 | SLC35A2 | NM_005660.3:c.168C>G:p.Tyr56* | Posterior superior frontal gyrus | 2.55% | 2.6% | validated |
| 49 | PTPN11 | NM_002834.5:c.50A>G:p.Glu17Gly | Blood | 0.00% | 0.0% | refuted |
| 49 | PTPN11 | NM_002834.5:c.50A>G:p.Glu17Gly | Left anterior temporal lobe | 0.00% | 0.1% | refuted |
| 49 | PTPN11 | NM_002834.5:c.50A>G:p.Glu17Gly | Left inferior frontal gyrus | 7.34% | 5.4% | validated |
| 49 | PTPN11 | NM_002834.5:c.50A>G:p.Glu17Gly | Left parietal operculum | 0.55% | 0.5% | validated |
| 49 | ROCK2 | NM_004850.5:c.4046C>T:p.Pro1349Leu | Blood | 0.00% | 0.01% | refuted |
| 49 | ROCK2 | NM_004850.5:c.4046C>T:p.Pro1349Leu | Left inferior frontal gyrus | 0.80% | 0.51% | validated |
| 49 | ROCK2 | NM_004850.5:c.4046C>T:p.Pro1349Leu | Left parietal operculum | 2.29% | 3.53% | validated |
| 49 | ROCK2 | NM_004850.5:c.4046C>T:p.Pro1349Leu | Left anterior temporal lobe | 0.00% | 0.69% | validated |
