## Supplemental Table 4 for "Detection of brain somatic variation in epilepsy-associated developmental lesions"

| **Patient** | **Gene name** | **Orientation** | **Primer sequence (5' -> 3')** |
| --- | --- | --- | --- |
| 11 | SLC35A2 | Forward | AGCCTGAGCTGCCTTTG |
|  |  | Reverse | CGGCCACTGGATCAGAAC |
|  | VEGFC | Forward | CAATCTTAGCTCATTTGTGGTCTTT |
|  |  | Reverse | CATGTACGAACCGCCAGAA |
| 13 | SDHA | Forward | TGGTATGGCTGCTTCTATGGATTA |
|  |  | Reverse | TGCACAGAGCCCACTGTCTG |
| 18 | PTPN11 | Forward | CTTCGTAGGTGTTGACTGCGA |
|  |  | Reverse | GAGCCTGTCCTCCTGCTCAA |
| 20 | EEF2 | Forward | ACCTTGGGCAACAGGAAGTCAT |
|  |  | Reverse | TTGCAGATGATCACCATCCACCT |
| 22 | IGF1R | Forward | ACCCCAAGATGAGGCCTTCCTTCC |
|  |  | Reverse | CGGCTCGGGCAGCTTGTTCT |
| 23 | NAV2 | Forward | AGCAGCTCCACCCCTACTAA |
|  |  | Reverse | GCCTTTGAGCCCCCTTTACT |
| 24 | DYNC1H1 | Forward | CAGGCTCCTTCTATCATGTCAC |
|  |  | Reverse | GTGGAACAGGTTGGCATAGT |
| 28 | ADAM22 | Forward | GAGCGAGGGAAACGGACTC |
|  |  | Reverse | GCACAGAGGACGGCTAACTT |
|  | RYR2 | Forward | CGGAGACGGATGAGAATGAAAC |
|  |  | Reverse | TGAAGTCGGGTATCGTTGGG |
|  | APC2 | Forward | AGTGAACCTTGCAGCGGGCA |
|  |  | Reverse | TGCAGGCTGAACTGGGTGGC |
|  | GABRB3 | Forward | GCAGAGTAATATTTCACTCAGTGTT |
|  |  | Reverse | GGTCCAGGATCGTGTTTCCA |
|  | TARDBP | Forward | GGTGGTGGGATGAACTTTGG |
|  |  | Reverse | GCATGTTGCCTTGGTTTTGG |
|  | CCDC88A | Forward | CCAGTTCCCAGCCAAGATGTA |
|  |  | Reverse | TCTCTCCCTAATCCTTGCCAATAA |
| 29 | MTOR | Forward | TGTTGCCATTTCAGGGTTTCTG |
|  |  | Reverse | GAAGATCTGCGCCAGGATGA |
| 30 | KIF22 | Forward | AATTCGAATGGGAGAGGCGG |
|  |  | Reverse | GGACCACACTCCGACACATA |
| 34 | KMT2C | Forward | GTTGCTGGTGGCTGAGAGTA |
|  |  | Reverse | CCGGCCTACTACTGTTGACC |
| 36 | POLR1A | Forward | CACGGCAAACACATCCTTGAT |
|  |  | Reverse | GCCACCTCTTGACCACTTTGT |
| 38 | PTPN11  SLC26A1 | Forward | CTTCGTAGGTGTTGACTGCGA |
|  |  | Reverse | GAGCCTGTCCTCCTGCTCAA |
| 41 |  | Forward | TCGTGCACACTGAGGAACA |
|  |  | Reverse | CTTCCACACAGTGGTCATCG |
| 42 | RAI1 | Forward | CCCCAAAAAGACAACTGGTCC |
|  |  | Reverse | ACAAGCATCCCCCAGGTTTTC |
| 44 | ENPP1 | Forward | CTCATTTTCGTTCTTCAGGACAGT |
|  |  | Reverse | TTACGTGGTGGGGAGAGGAAC |
| 45 | PTPN11 | Forward | GACTGCGATATTGACGTTCCC |
|  |  | Reverse | TGAGAATCCGCATGCCAGC |
|  | BRAF | Forward | GCAGCATCTCAGGGCCAAAA |
|  |  | Reverse | TGCTTGCTCTGATAGGAAAATGAG |
| 47 | RHEB | Forward | CTCCTGCACTCAAAGCCTCC |
|  |  | Reverse | GCCAACACACACTAAGCTCTTG |
| 48 | SLC35A2 | Forward | TTGGCTTCCTTTCTGTCGGG |
|  |  | Reverse | TGAAGTGGGTGGGTTCATGG |
| 49 | PTPN11 | Forward | ATGCGTCTTAGCTGTGTTGT |
|  |  | Reverse | ACGGAAAGTGTGAAGTCTCCAG |
|  | ROCK2 | Forward | GTTTGTTTGGGGCAAGCTGT |
|  |  | Reverse | GAAGAGCAGCAGAAGTGGGT |
